# Environmental monitoring shows SARS-CoV-2 contamination of surfaces in food plants

**DOI:** 10.1101/2020.12.10.20247171

**Authors:** Ziwen Ming, Sukkyun Han, Kai Deng, Youngsil Ha, SungSoo Kim, Enrique Reyes, Yu Zhao, Anatoly Dobritsa, Meiting Wu, Dandan Zhang, David P Cox, Emma Joyner, Hemantha Kulasekara, Seong Hong Kim, Yong Seog Jang, Curtis Fowler, Xing Fei, Hikari Akasaki, Eni Themeli, Alexander Agapov, Dylan Bruneau, Thao Tran, Cameron Szczesny, Casey Kienzle, Kristina Tenney, Hao Geng, Mansour Samadpour

## Abstract

The SARS-CoV-2 pandemic has presented new challenges to food manufacturers. In addition to preventing the spread of microbial contamination of food, with SARS-CoV-2, there is an additional focus on preventing SARS-CoV-2 infections in food plant personnel. During the early phase of the pandemic, several large outbreaks of Covid-19 occurred in food manufacturing plants resulting in deaths and economic loss. In March of 2020, we assisted in implementation of environmental monitoring programs for SARS-CoV-2 in 116 food production facilities. All participating facilities had already implemented measures to prevent symptomatic personnel from coming to work. During the study period, from March 17, 2020 to September 3, 2020, 1.23% of the 22,643 environmental samples tested positive for SARS-CoV-2, suggesting that infected individuals are actively shedding virus. Virus contamination was commonly found on frequently touched surfaces. Most plants managed to control their environmental contamination when they became aware of the positive findings. Comparisons of the personnel test results to environmental contamination in one plant showed a good correlation between the two. Our work illustrates that environmental monitoring for SARS-CoV-2 can be used as a surrogate for identifying the presence of asymptomatic and pre-symptomatic personnel in workplaces and may aid in controlling infection spread.

**Highlights:**

- Environmental contamination by SARS-CoV-2 virus was detected in food plants
- Out of 22,643 environmental swabs, 278 (1.23%) were positive for SARS-CoV-2
- Frequently touched surfaces had the most contamination
- Surface testing for SARS-CoV-2 may indicate presence of asymptomatic carriers

## Introduction

Severe acute respiratory syndrome coronavirus 2 (SARS-CoV-2), a highly infectious novel coronavirus, which originated from a food market in Wuhan, China, in December 2019, has caused a global pandemic (*3*). As of November 1st, 2020, there have been nearly 46 million confirmed cases of coronavirus disease 2019 (COVID-19), with 1.2 million deaths globally (*12*).

The transmission of COVID-19 is facilitated mainly through direct personal contact and respiratory droplets (*1*). Additionally, contaminated surfaces were also reported as another mode of transmission (*5, 6*). SARS-CoV-2 remains viable on surfaces for days (*9*). The virus is reported to be stable on surfaces such as metal, glass and plastic for up to 9 days (*4*). Surface disinfection with 62-71% ethanol, 0.5% hydrogen peroxide or 0.1% sodium hypochlorite can efficiently inactive the virus within 1 minute (*4*).

During the early phase of the pandemic, high SARS-CoV-2 positive rates among environmental surfaces were associated with a large number of COVID-19 cases in places such as hospitals and living spaces (*13*). At one hospital in Italy, air and surfaces had the most positives within the areas designated for patients (*7*). In a study of 112 surface samples taken from the living quarters of 13 laboratory-confirmed COVID-19 cases, Wei et al. (*11*) found that 44 (39.3%) of the samples were positive for SARS-CoV-2 RNA. Research on built environments emphasized the importance of proper disinfection of toilet areas, sanitization of surfaces, open space, and window ventilation which can effectively limit the concentration of SARS-CoV-2 (6)

According to the US Centers for Disease Control and Prevention, USA (CDC), of the 130,578 people employed in the food industry who were tested for COVID-19 in April, 3% tested positive (*2*). In a more comprehensive analysis, the CDC analyzed Covid-19 infections in the food industry in 30 states spanning a period from March to May of 2020. Results showed that some food plants had infection rates as high as 43%. Most workers who were Covid-19 positive were ethnic minorities (83.2%). The asymptomatic rate was about 15%, indicating that screening for Covid-19 symptoms alone is not adequate to combat infection spread (*10*). Although it is highly likely that airborne transmission substantially contributed to observed infection outcomes in the CDC study, transmission could have additionally occurred through SARS-CoV-2 contaminated surfaces (*8*). However, the extent of virus contamination in the food industry has not been reported yet. Here we present the analysis of 22,643 surface samples from food processing facilities in the USA, tested for the presence of SARS-CoV-2 RNA. Such data can help food manufacturing plants to evaluate their decontamination protocols and can be used as a surveillance method for detecting viral shedding from asymptomatic/presymptomatic infections among workers.

## MATERIALS AND METHODS

### Environmental Sampling

IEH SARS-CoV-2 Surface Swab Kits (P/N: PS-S02; Microbiologique Inc., Lake City Way NE, WA) were used for sampling environmental surfaces. Sampling was performed according to the manufacturer’s instruction by plant personnel. The sampling buffer impregnated swab was applied to designated areas, which were thoroughly swabbed before the swab was transferred to a transport vial containing viral transport medium (VTM). The vials were packed, labeled, and shipped to Molecular Epidemiology Inc. (Lake Forest Park, WA, USA) for testing.

### RNA Extraction

Total nucleic acids were extracted and purified from environmental swabs using the IEH Nucleic Acid Extraction Reagent Kit (P/N: PM-23; Microbiologique Inc., Seattle, WA, USA) in an automated KingFisher-96 (Thermo Fisher Scientific Inc., Waltham, MA, USA) nucleic acid purification system. MS2–RPP, an engineered MS2 phage particle encapsulating an RNA fragment from the human ribonuclease P gene (RNase P), served as the extraction control for RNA extraction and was processed with every set of samples.

### RT-PCR

The IEH SARS-CoV-2 RT-PCR Test kit (P/N: PM-22; Microbiologique, Seattle, WA, USA) was used for the detection of RNA from SARS-CoV-2 in environmental samples. This test kit was derived from the SARS-CoV-2 diagnostic test *“CDC 2019-Novel Coronavirus (2019-nCoV) RT-PCR Diagnostic Panel”*, developed by the CDC. This kit contains N1, N2 and RPP primer/probe sets to detect the viral nucleocapsid gene and the human RNase P RNA (internal control).

### Clinical specimen analysis

Clinical specimens from Establishment E were processed at the CLIA certified laboratory of Molecular Epidemiology Inc. (Lake Forest Park, WA, USA) using the same RNA extraction method and the RT-PCR method and using N1/RPP and N2 primers/probes.

### Data analysis

The cycle threshold (Ct) values from the RT-PCR results were used as indicators of the copy number of SARS-CoV-2 RNA in environmental samples with lower cycle threshold values corresponding to higher viral copy numbers. Descriptive statistical analyses were performed for both environmental data and clinical data by using SigmaPlot 14.0 (Systat Software, Inc).

## RESULTS

A total of 22,643 environmental samples with source information were received between March 17th, 2020 and September 3rd, 2020 from 116 food manufacturing plants. Of these, 278 environmental samples (1.23%) were found to be positive for SARS-CoV-2 during the study period (Table 1). Samples were analyzed by grouping into workplace source and surface types. Workplace sources were divided into three groups: welfare area, working area, and entrance. The welfare area and working areas had swabs that came from distinctly different locations based on usage and were thus subdivided. The welfare area was split into three categories, which were bathrooms, lunchrooms, and locker rooms. The working area comprised of office/conference rooms, training rooms, and receiving rooms. The entrance and other ingress points had the highest occurrence of positive SARS-CoV-2 swabs among tested areas, with a frequency of 1.57% (38/2439) (Table 1). There were 32 out of 2255 (1.42%) positive samples reported in the working area group, with 25/1883 (1.33%) positive from office/conference rooms and 1/265 (0.38%) positive from training rooms. The receiving room showed the highest positivity rate of 5.61% (6/107). An overall total of 1.36% (139/10226) of samples were positive in the welfare area, where bathrooms, lunchrooms, and locker rooms tested positive with percentages of 1.17% (31/2640), 1.33% (70/5260), and 1.63% (38/2326), respectively.

**Table 1.** Results summary of environmental testing for SARS-CoV-2, in different work areas in food plants.

| Sites | Positive tests | Total tests | Percent Positive (%) |
| --- | --- | --- | --- |
| **Welfare Area** |  |  |  |
| Bathroom | 31 | 2640 | 1.17 |
| Lunchroom | 70 | 5260 | 1.33 |
| Locker room | 38 | 2326 | 1.63 |
| **Subtotal** | 139 | 10226 <sup>a</sup> | 1.36 |
| **Working area** |  |  |  |
| Office/conference | 25 | 1883 | 1.33 |
| Receiving | 6 | 107 | 5.61 |
| Training | 1 | 265 | 0.38 |
| **Subtotal** | 32 | 2255 <sup>a</sup> | 1.42 |
| **Building Entrance/Hall** | 38 | 2439 | 1.56 |
| **Not-specified <sup>b</sup>** | 69 | 7723 | 0.89 |
| **Total** | 278 | 22643 | 1.23 |

<sup>a</sup> Total number of tests done for each area group.
<sup>b</sup> Results from surface samples in which area information was lacking.

We also analyzed what surface types were more abundant with the virus (summarized in Table 2). Among the positive samples, 46 (16.55%) had no specified surface information, and 93 (33.45%) were found on doorknobs/handles, the surface with the highest positive frequency rate. Other surfaces that frequently tested positive for SARS-CoV-2 included tables/counters (21), computer devices (20), sanitizer-dispensers (17), switches (12), rails (12), chairs/benches (11) and timeclocks (10). There were 36 positive samples found on other surfaces.

**Table 2.** Results summary for environmental testing for SARS-CoV-2 in different surface areas.

| Surface | Area <sup>a</sup> | Total Positive<br>Samples (n=278) | Percent Positive (%) |
| --- | --- | --- | --- |
| Computer devices |  | 20 | 7.19 |
|  | Office | 20 |  |
| Chair/bench |  | 11 | 3.96 |
|  | breakroom | 1 |  |
|  | Locker Room | 7 |  |
|  | Not-specified <sup>b</sup> | 3 |  |
| Sanitizer Dispensers |  | 17 | 6.12 |
|  | bathroom | 2 |  |
|  | breakroom | 6 |  |
|  | Entrance/hallway | 2 |  |
|  | Locker Room | 4 |  |
|  | Not-specified | 3 |  |
| Door knob/ handles <sup>c</sup> |  | 93 | 33.45 |
|  | bathroom | 19 |  |
|  | breakroom | 15 |  |
|  | Entrance/hallway | 18 |  |
|  | Locker Room | 8 |  |
|  | Office | 5 |  |
|  | Not-specified | 28 |  |
| Rail |  | 12 | 4.32 |
|  | breakroom | 1 |  |
|  | Entrance/hallway | 3 |  |
|  | Not-specified | 8 |  |
| **Switch** |  | 12 | 4.32 |
|  | bathroom | 3 |  |
|  | breakroom | 3 |  |
|  | Locker Room | 1 |  |
|  | Office | 1 |  |
|  | Not-specified | 4 |  |
| **Table/Counter** |  | 21 | 7.55 |
|  | breakroom | 13 |  |
|  | Locker Room | 1 |  |
|  | Office | 2 |  |
|  | Not-specified | 5 |  |
| **Timeclock** |  | 10 | 3.6 |
|  | Entrance/hallway | 3 |  |
|  | Not-specified | 7 |  |
| **Others <sup>d</sup>** |  | 36 | 12.95 |
| **Not-specified <sup>e</sup>** |  | 46 | 16.55 |

<sup>a</sup> Work areas belonging to specific surface types.
<sup>b</sup> Results from surface samples in which area information was lacking
<sup>c</sup> Including doorknobs, door pushbars, handles of devices such as microwave, refrigerator, faucet and etc.
<sup>d</sup> Number of positive test persurface that is less than ten were combined in this group.
<sup>e</sup> Surface is not indicated by customers, only area information is provided.

**Figure 1.**
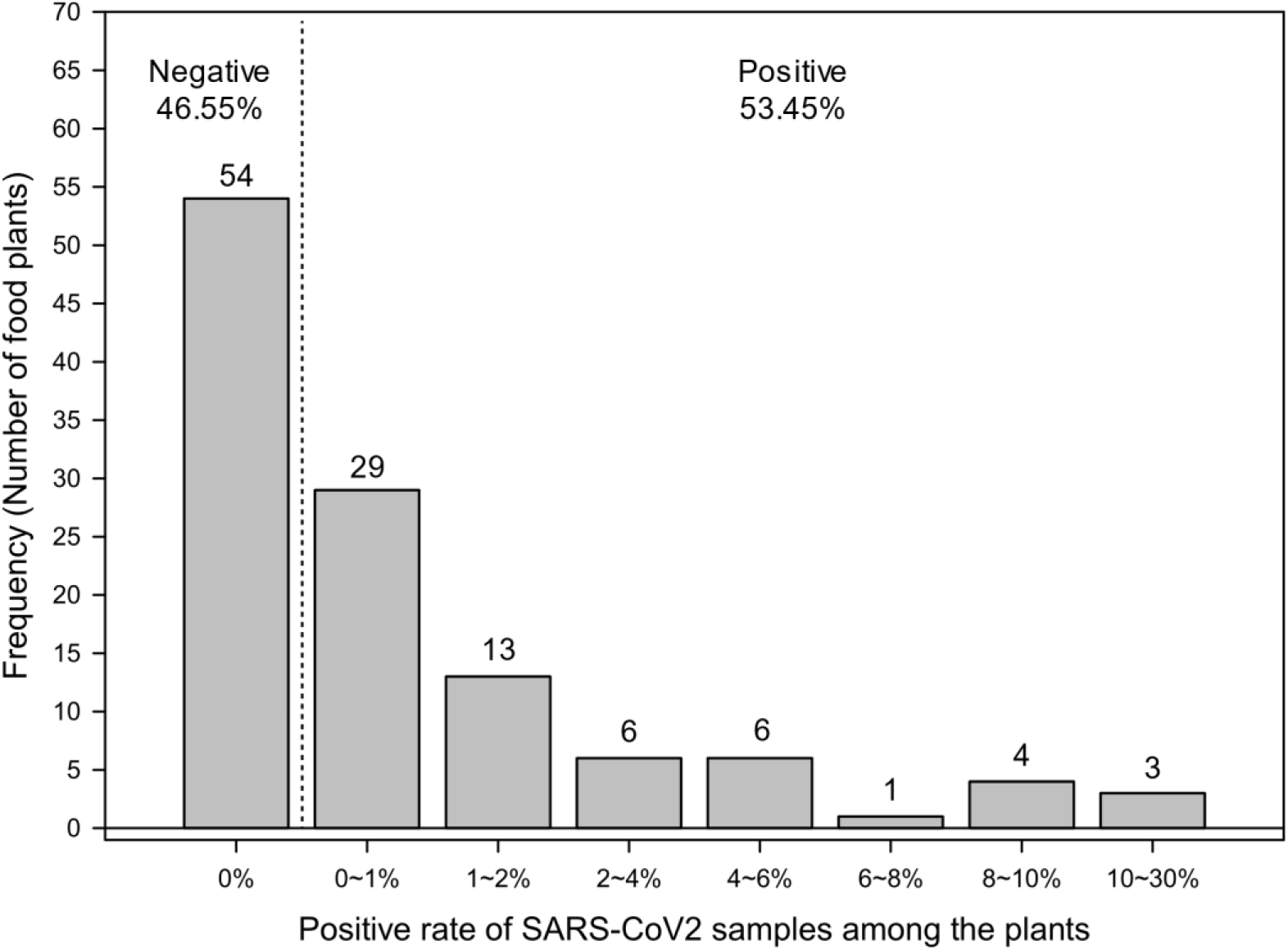
Histogram of percentage of positive cases for each individual plant (n=116). The upper bounds of each interval ranges are inclusive while lower bounds are excluded. 0% has no interval range but only containing one value.

We next analyzed to what extent these 116 food processing plants were affected. Figure 2 shows the frequency distribution of the percentage of positive cases for each individual plant. A total of 53% (62/116) of food plants had at least one positive sample through the observational period. The positive rate of individual plants ranged from 0% to 30% with a median level of 0.25%.

**Figure 2.**
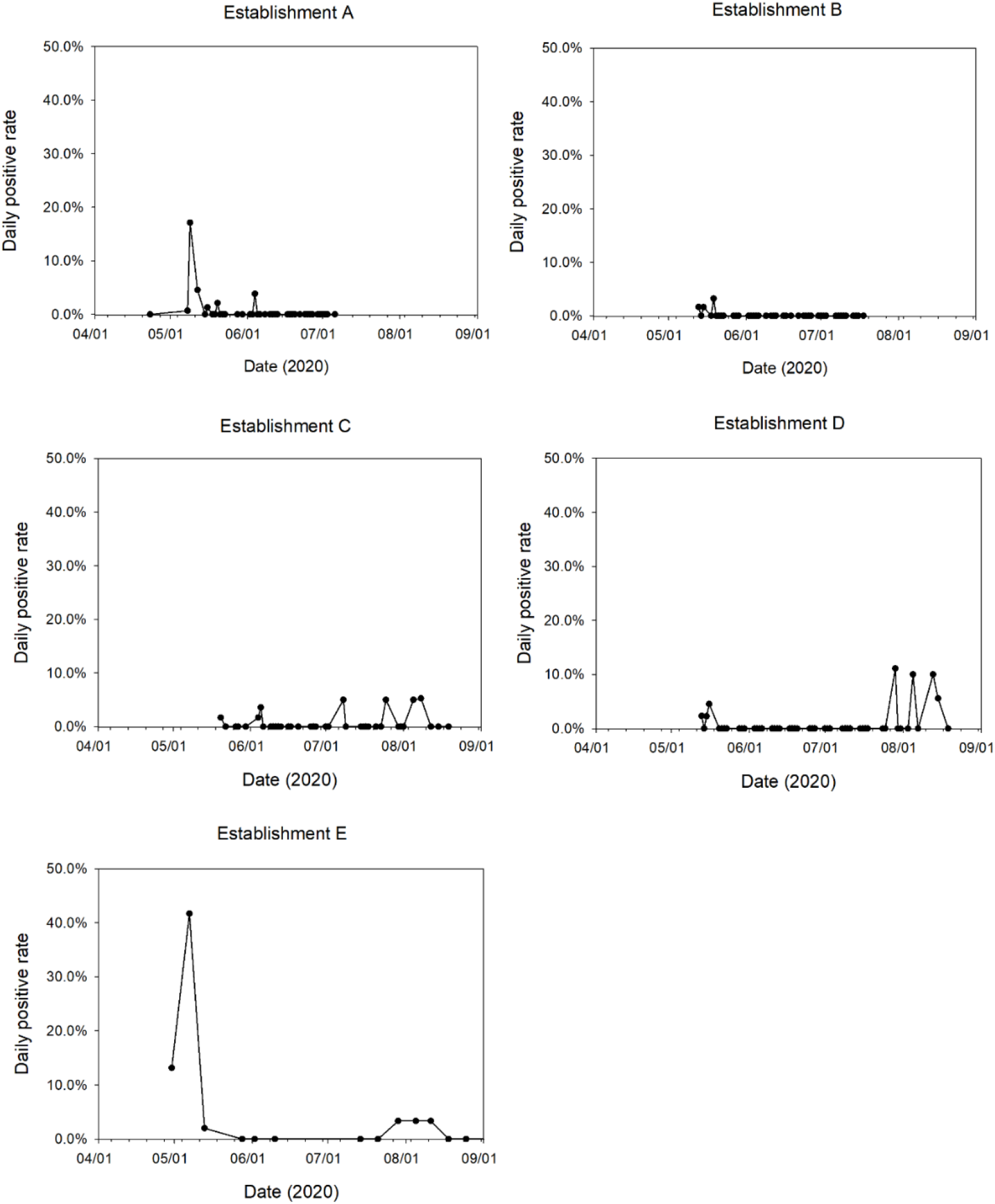
Positive environmental samples in five food establishments during the study period (Daily positive rate = number of daily positive samples / number of daily total samples ×100%).

As the time progressed from the early phase of the pandemic in February, almost all food manufacturing companies implemented a variety of safety measures such as vigorous decontamination, mandatory personal protective equipment, symptom screening, and SARS-CoV-2 screening. We therefore undertook a longitudinal observational study of five companies for timeline data analysis to determine the outcomes of SARS-CoV-2 preventive measures (Figure 2). During the study period (March 17^th^-September 3^rd,^ 2020), 1477, 1577, 912, 867 and 433 surface specimens were received from establishments A, B, C, D and E, respectively. A decreasing trend of daily positive rate was observed in establishments A, B and E after reaching peaks on 5/9/2020, 5/19/2020 and 5/7/2020. The other two establishments continued to have sporadic findings of environmental contamination with the virus.

Establishment E additionally submitted 1248 human nasopharyngeal specimens from plant personnel for SARS-CoV-2 diagnosis from May 4th to June 18th, 2020. Our results indicated that 10.90% of human samples were positive for SARS-CoV-2 (Figure 3). This indicated that even with screening for symptomatic individuals and strict environmental decontaminations, asymptomatic and pre-symptomatic individuals presumably contaminated their surroundings with the virus.

**Figure 3.**
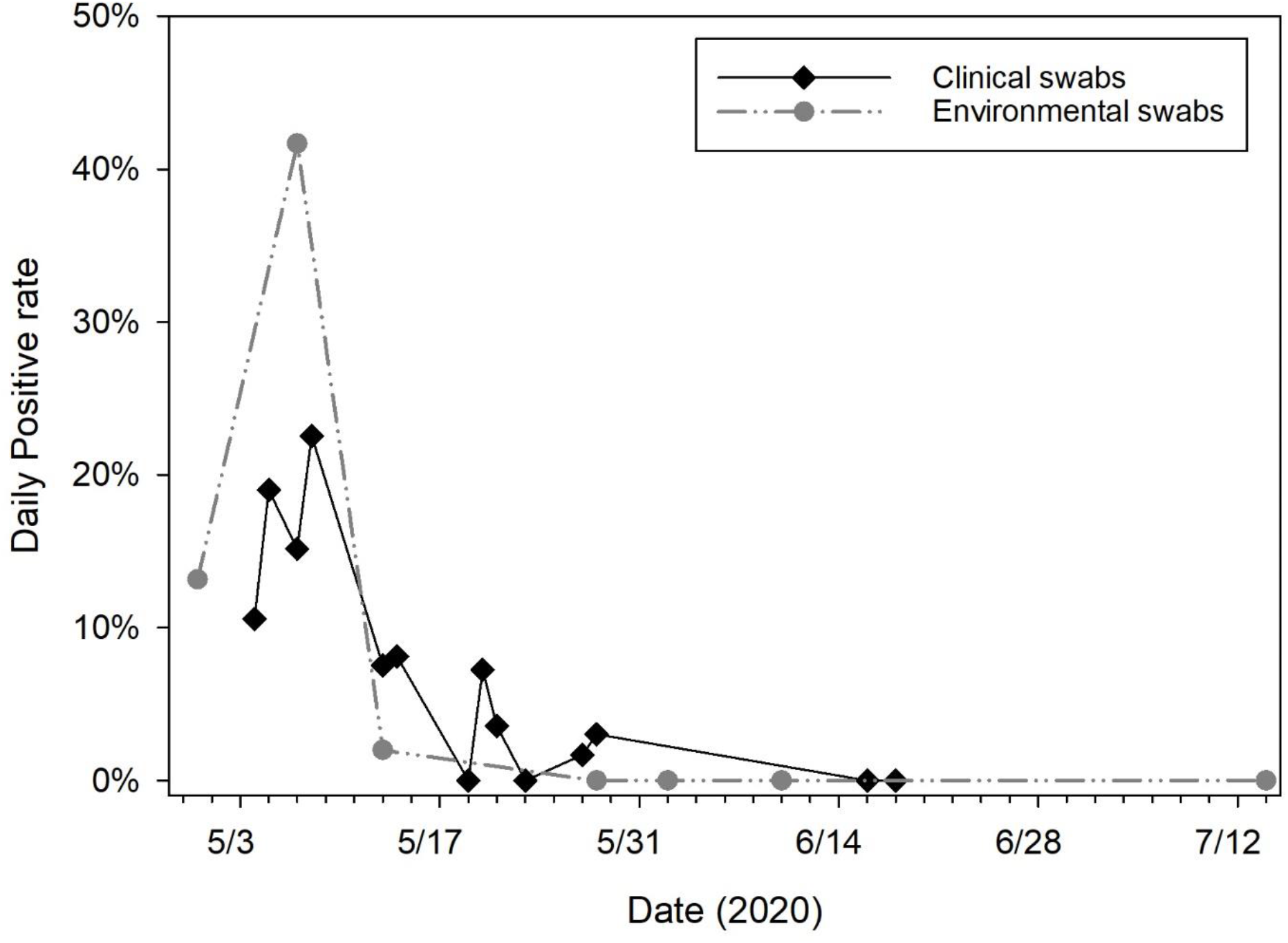
Daily Positive rate of Clinical samples vs. Environmental samples of Establishment E from April 30 to July 18, 2020.

## DISCUSSION

The goal of our study was to provide a tool for monitoring the presence of SARS-CoV-2 in food production facilities. Environmental monitoring for foodborne pathogens such as *Listeria* and *Salmonella* are routinely conducted in food production facilities to document the sanitary conditions under which food is produced. The environmental monitoring results provide a constant feedback to the sanitation and food safety teams, based on which they can take corrective actions. The SARS-CoV-2 pandemic has presented a new challenge to food manufacturers. While food production is focused on preventing the spread of microbial contamination of foods, with SARS-CoV-2, there is an additional focus on operating while protecting personnel from the SARS-CoV-2 virus. Early on, consensus were developed around using personal protective equipment (face masks, face shields, gloves, plastic and Plexiglas barriers), health monitoring of the personnel, reducing density in otherwise crowded areas, frequent sanitation of surfaces, closing down or reducing the capacity in break rooms, contact tracing, quarantine of exposed personnel, and testing.

Currently there is not much information available about the level of work-place contamination due to asymptomatic and pre-symptomatic COVID-19 infections. In March of 2020 we assisted in implementation of environmental monitoring programs for SARS-CoV-2 in several food production facilities. All facilities had implemented measures to prevent symptomatic/pre-symptomatic personnel from coming to work. During the study period 278 of the 22,643 samples (1.23%) tested positive for SARS-CoV-2. 62 of the 116 food production facilities had at least one positive sample for SARS-CoV-2. Among the production facilities the rate of positive samples ranged from 0-30%.

The data (Table 1 and 2) clearly show that all frequently touched areas can be expected to have contamination. Door knobs/handles had the highest rate of contamination (33.5%), followed by computers, desks/tables, sanitizer dispensers, hand rails, switches, chairs/benches, and timeclocks. Figure 2 shows the course of contamination in five plants over the study period. As environmental contamination with SARS-CoV-2 is a reflection of the infection among personnel, it is to be expected that contamination would be detected sporadically over time. In establishments A and B contamination was detected early on, and all subsequent samples were negative for SARS-CoV-2 virus. While in the other establishments the viral contamination appears and disappears over time. Although a positive SARS-CoV-2 RT-PCR test of an environmental sample does not necessarily mean that the sampling site is contaminated with infectious viral particles, it is a clear indication of active shedding of the virus by infected individuals. SARS-CoV-2 contamination occurred even with decontamination protocols implemented in place, indicating that they were inadequate. During the course of this study, our findings enabled all participating production facilities to fine-tune their COVID safety protocols and helped them make decisions regarding personnel testing.

In Establishment E where we performed environmental testing, initial high positive rates (∼40%) prompted testing of personnel. Comparisons of the human and environmental samples taken in the same facility at the same time showed that 10.90% of human and 8.54% of the environmental samples were positive for the virus. This shows that in the absence of personnel testing, environmental testing for SARS-CoV-2 could indicate active human infections.

Our results clearly show that monitoring for the SARS-CoV-19 in work places can be a valuable tool in the control of the spread of SARS-CoV-2 virus. The limitation of the study is that we were not able to determine the role of the environmental contamination in the spread of the infection. This is because RT-PCR test cannot differentiate between infectious and non-infectious virus.

## Data Availability

Unprocessed data used in the manuscript is available upon request.

